# Vitamin D supplementation to prevent tuberculosis infection and disease: systematic review and individual participant data meta-analysis of randomised controlled trials

**DOI:** 10.64898/2026.11.04.26367248

**Authors:** David A Jolliffe, Keren Middelkoop, Davaasambuu Ganmaa, Boldbaatar Irmuun, John D Sluyter, Christopher R Sudfeld, Wafaie W Fawzi, Adrian R Martineau

**Affiliations:** Queen Mary and Barts Health Tuberculosis Centre, Blizard Institute, Faculty of Medicine and Dentistry, Queen Mary University of London, London, UK; Desmond Tutu HIV Centre, Institute of Infectious Disease and Molecular Medicine, University of Cape Town, Cape Town, South Africa; Department of Nutrition and Department of Global Health and Population, Harvard T.H. Chan School of Public Health, Boston, MA, USA; Mongolian Health Initiative (MHI), Ulaanbaatar, Mongolia; School of Population Health, University of Auckland, Auckland, New Zealand

**Keywords:** vitamin D, tuberculosis, tuberculosis infection, randomised controlled trials, individual participant data meta-analysis

## Abstract

**Background:** Several randomised controlled trials of vitamin D supplementation for prevention of tuberculosis infection and disease have been conducted, but no individual participant data (IPD) meta-analysis has been conducted.

**Methods:** We conducted a systematic review, one-stage and two-stage IPD meta-analysis (PROSPERO CRD420251173446) of double-blind, placebo-controlled randomised trials of vitamin D supplementation for prevention of tuberculosis infection or disease. We searched MEDLINE, Embase, Scopus, Cochrane and ClinicalTrials.gov from inception to October 29, 2026. The co-primary outcomes were incident tuberculosis infection (evidenced by tuberculin skin test or Interferon Gamma Release Assay conversion) and incident tuberculosis disease. Mixed-effects models estimated adjusted odds ratios (aORs), accounting for trial and prespecified covariates. Prespecified subgroup analyses investigated effect modification by sex and age, body-mass index, vitamin D status, and BCG vaccination status.

**Findings:** Four trials contributed IPD for 14,619 participants. Vitamin D did not reduce risk of incident tuberculosis infection (9,557 participants; aOR 0.97, 95% CI 0.81-1.18; p=0.79) or incident tuberculosis disease (14,533 participants; aOR 0.78, 95% CI 0.56-1.10; p=0.15). There was no evidence of effect modification by participant characteristics investigated in sub-group analyses, including in participants with 25-hydroxyvitamin D <25 nmol/L (infection: aOR 0.95, 95% CI 0.67-1.34; disease: 1.01, 0.45-2.30). Serious adverse events and mortality were unaffected.

**Interpretation:** Vitamin D supplementation did not reduce the risk of incident tuberculosis infection or disease in the populations studied, either overall or within sub-groups. The 95% confidence interval of the effect estimate for prevention of tuberculosis disease remains compatible with a potentially important benefit, and further trials for this indication may be warranted.

## Research in Context

### Evidence before this study

Before undertaking this study, we conducted a scoping search for systematic reviews, meta-analyses, and randomised controlled trials evaluating vitamin D supplementation for prevention of tuberculosis infection or disease. We subsequently searched MEDLINE, Embase, Scopus, the Cochrane Central Register of Controlled Trials and ClinicalTrials.gov from database inception to October 29, 2026, without language restrictions, using the general search terms ‘vitamin D’, ‘tuberculosis’, ‘prevention’ and ‘randomised controlled trial’, to identify primary trials eligible for this systematic review and individual participant data meta-analysis. The scoping search identified two previous aggregate-data meta-analyses. One combined incident tuberculosis infection and tuberculosis disease into a single preventive outcome (risk ratio 0.75, 95% CI 0.56–1.01), despite their distinct biological and clinical meanings. The other reported no significant effect on tuberculosis infection (odds ratio 0.95, 95% CI 0.79–1.14) or disease (0.77, 0.56–1.05), with moderate certainty, but included Dude et al. (2022) and Middelkoop et al. (2023) as independent trials, although the reported populations had identical mean age, sex distribution, and baseline 25-hydroxyvitamin D concentration, while being assigned different sample sizes, dosing regimens, and countries. The effect estimates attributed to Dude et al. could not be linked to an independently identifiable eligible trial, raising substantial uncertainty about whether it represented an independent dataset; counting it separately might therefore have duplicated trial evidence and overstated the precision of the pooled estimates. Furthermore, neither review used individual participant data and therefore could not harmonise outcome definitions and analysis populations, verify participant eligibility, adjust consistently for baseline covariates, or reliably investigate participant-level effect modification, including by baseline vitamin D status.

### Added value of this study

This study is the first IPD meta-analysis of randomised trials evaluating vitamin D supplementation for prevention of tuberculosis infection and disease. We obtained IPD from all four eligible trials, comprising 14,619 participants, and applied harmonised outcome definitions and one-stage mixed-effects models. Access to IPD allowed us to examine, using definitions applied consistently across trials, whether the effect of vitamin D differed by participant characteristics, including among people with profound vitamin D deficiency (baseline 25-hydroxyvitamin D <25 nmol/L), in whom benefit is most plausible. Vitamin D supplementation did not significantly reduce incident tuberculosis infection (evidenced by tuberculin skin test or Interferon Gamma Release Assay conversion) or incident tuberculosis disease, and there was no evidence of effect modification by sex or baseline age, body-mass index, vitamin D status or BCG vaccination status. The 95% CI of the estimate for prevention of tuberculosis disease was wide and remained compatible with a potentially clinically important reduction in risk.

### Implications of all the available evidence

Taken together, the available evidence does not support routine vitamin D supplementation to prevent tuberculosis infection or disease in children or adults. The relatively precise null estimate for tuberculosis infection, and no evidence of subgroup benefit, makes a large overall preventive effect unlikely, although a modest reduction of up to about 19% in the odds of infection cannot be ruled out. For tuberculosis disease, uncertainty means that a clinically important benefit cannot be fully excluded; there was no evidence of effect modification by participant characteristics, including baseline vitamin D status, although power for subgroup analyses of tuberculosis disease was limited.

## Introduction

Active tuberculosis (TB) continues to impose a devastating global burden, with an estimated 10.7 million new cases and 1.23 million deaths in 2024.^1^ Most TB arises from progression of asymptomatic tuberculosis infection (TBI), which is highly prevalent: around 23% of the global population is sensitised to *M. tuberculosis* (Mtb),^2^ with 3.7% estimated to be harbouring viable Mtb.^3^ Since TBI carries a 5-10% lifetime risk of progression in HIV-uninfected people^4^ – and a similar annual risk in people living with HIV^5^ – this represents a vast reservoir for future disease reactivation.^2^ Vaccination and tuberculosis preventive treatment (TPT) have limited efficacy in reducing risk of progression to active TB in Mtb-sensitised individuals,^6,7^ and effective interventions to prevent TBI are lacking.^8,9^ There is therefore an urgent need for novel, cost-effective interventions that can induce protection against acquisition of TBI and reduce the risk of progression to active TB in Mtb-infected people.

Vitamin D has emerged as a candidate for prevention of both TBI and TB disease, primarily due to its role in enhancing innate antimicrobial effector responses including production of reactive oxygen and nitrogen intermediates,^10,11^ antimicrobial peptides^12,13^ and induction of autophagy. Epidemiological evidence supports this link: numerous observational studies report independent associations between low vitamin D status and increased risk of TBI and/or progression to active TB.^14,15^

Results from randomised controlled trials (RCTs) of vitamin D supplementation for prevention of TBI and TB disease are mostly null,^16–19^ but previous meta-analyses have been on aggregated data, have either combined TBI and TB disease^20^ or included potentially non-independent trial evidence,^21^ and could not reliably investigate if the effect of vitamin D on outcomes differs by participant-level characteristics. By harmonising the analysis of individual participant data (IPD) from 4 RCTs (n=14,653 participants) we sought to provide a definitive answer to the question of vitamin D’s protective efficacy on risk of incident TBI or TB disease, overall and in sub-groups stratified by participant-level age, sex, baseline vitamin D status, body mass index (BMI) and BCG vaccination status.

## Methods

### Search strategy and selection criteria

The methods for this systematic review and meta-analysis were described in an online protocol that was registered with the PROSPERO International Prospective Register of Systematic Reviews, number CRD420251173446. Research ethics committee approval to contribute individual participant data (IPD) to the current meta-analysis was not required for any study. All contributing trials were approved by the relevant research ethics committees, and written informed consent was obtained from participants or their parents or guardians. Findings are reported according to the PRISMA guidelines for IPD meta-analysis.^22^

We included double-blind, placebo-controlled randomised controlled trials comparing oral vitamin D supplementation with placebo for the primary prevention of TBI, TB disease, or both. Eligible trials assessed incident TBI by conversion of a tuberculin skin test (TST) or interferon-gamma release assay (IGRA) from negative to positive, or incident TB disease according to trial-specific diagnostic criteria. We excluded non-randomised or non-placebo-controlled studies, trials evaluating vitamin D as treatment for established TBI or TB disease, and trials that did not report either outcome. For analyses of incident TBI and TB disease, we additionally excluded participants with the corresponding outcome present at baseline.

DAJ searched MEDLINE, EMBASE, Scopus, the Cochrane Central Register of Controlled Trials (CENTRAL) electronic databases and the ClinicalTrials.gov registry using search strategies described in the Supplementary Appendix. Searches were regularly updated up to and including 29th October 2026. Records from all sources were de-duplicated. Two investigators (DAJ and ARM) reviewed the search results and determined which studies met eligibility criteria.

### Data analysis

IPD were requested from the Principal Investigator for each eligible trial, and the terms of collaboration were specified in a data transfer agreement, signed by representatives of the data provider and the recipient (Queen Mary University of London). Data were de-identified at source prior to transfer via email in password-protected files. On receipt, DAJ assessed data integrity by performing internal consistency checks and by cross-checking proportions of participants with incident TBI and/or TB disease against those reported in published papers. Study authors were contacted to provide missing data and to resolve queries arising from these integrity checks. Once queries had been resolved, clean data were uploaded to the main study database, which was held in STATA v19 (College Station, TX, USA).

Data relating to study-level characteristics were collected, including study setting, participant eligibility, details of the intervention and control regimens (dose, frequency, and duration), and duration of follow-up. Individual participant data (IPD) were extracted for the following baseline characteristics: age, sex, BMI, HIV status, BCG vaccination status, and study allocation (vitamin D vs. placebo). Baseline circulating 25(OH)D concentrations were recorded, along with the specific assay used (chemiluminescence immunoassay, enzyme immunoassay, enzyme-linked fluorescent assay, or liquid chromatography–tandem mass spectrometry) and the associated quality assurance schemes. Follow-up data were collected for the occurrence of incident TBI, incident TB disease and secondary outcomes of severe adverse events, hypercalcaemia and renal stones. Seasonally adjusted 25(OH)D concentrations were used where available. For the Sudfeld et al. trial, concentrations were deseasonalised using an unadjusted 12-month cosinor model: observed 25(OH)D concentration was regressed on sine and cosine terms derived from the calendar month of blood sampling, and each participant’s model-predicted seasonal deviation from the annual mean was subtracted from their observed concentration. This provided an estimate of each participant’s annual mean 25(OH)D concentration.^23^ Seasonally adjusted values were supplied by investigators for the Ganmaa et al. 2020 and Middelkoop et al. 2023 trials; seasonal adjustment was not possible for Ganmaa et al. 2012 because dates of blood sampling were unavailable.

To evaluate risk of bias within studies, we used the Cochrane Collaboration Risk of Bias tool^24^ to assess the following variables: sequence generation, allocation concealment, blinding of participants, personnel and outcome assessors, completeness of outcome data, evidence of selective outcome reporting and other potential threats to validity. Selectivity of reporting was assessed either by comparing study protocols against study reports, or by specifically asking study authors whether all pre-specified outcomes were reported. Study quality was assessed independently by two investigators (ARM and DAJ). Discrepancies were resolved by consensus. The certainty of evidence for each outcome was assessed using the GRADE approach (Table S3).^25^

For risk of bias across studies, funnel plots and statistical tests for funnel plot asymmetry were not undertaken because only three trials contributed to each co-primary outcome analysis, making meaningful assessment of small-study effects unfeasible.

The co-primary outcomes of the meta-analysis were the proportion of participants acquiring incident TBI and the proportion of participants developing incident TB disease during the follow-up period. Incident TB infection was defined as a positive result from a TST or IGRA performed at follow-up in an individual who had a negative test result at baseline. Incident TB disease was defined according to diagnostic criteria of the trials, with individuals starting full TB treatment categorised as having active TB (Table S5). To ensure that analysis was restricted to truly incident cases, individuals with TB infection or active TB disease at baseline were excluded from the respective primary outcome analyses. Secondary outcomes included all-cause mortality, TB-specific mortality, and incidence of serious adverse events (SAEs) of any cause. Additionally, safety outcomes relating to recognised complications of vitamin D supplementation (adverse reactions) were defined as the occurrence of at least one episode of hypercalcemia or renal stones.

Data were analysed using STATA version 19. The co-primary analyses were independently replicated in R (version 4.6.1; lme4 and meta packages) by a second statistician (JDS). Our individual participant data (IPD) meta-analysis followed established guidelines,^26^ employing both 1-stage and 2-stage approaches to assess the co-primary outcome of incident TBI, and 1-stage only to assess the co-primary outcome of incident TB disease due to scarcity of events.

In the 1-stage approach, IPD from all included trials were modelled simultaneously using logistic regression models to produce pooled adjusted odds ratios (ORs), associated 95% confidence intervals (CIs), and p-values. These models were corrected for the fixed effects of a minimum set of mutually available factors (age, sex and trial). For the co-primary outcome of incident TBI, mixed models were used with additional adjustment for the effects of trial-specific stratification factors using a random intercept. For the co-primary outcome of incident TB disease, model convergence was not possible using this method due to the relatively low event rate. For this analysis, standard logistic regression models were used and a variance-covariance estimator was fitted to account for stratified randomisation, and singleton clusters were assigned to participants in one trial which did not stratify randomisation^17^ to prevent exclusion of their data. In the 2-stage approach for incident TBI, a separate analysis was first performed within each individual trial using the equivalent method and corrective factors to obtain aggregate data; these results were then combined using a standard meta-analysis model and presented in a forest plot. Between-study heterogeneity was assessed using Cochran’s Q test and quantified using the I^2^ statistic.

Pre-specified subgroup analyses were conducted within the one-stage framework to investigate effect modification by baseline age, sex, vitamin D status, BMI, and BCG vaccination status. Baseline 25(OH)D was dichotomised at 25 nmol/L and, separately, at 50 nmol/L. BMI categories were defined using age- and sex-standardised BMI-for-age z scores in children and absolute BMI values in adults: underweight was defined as a z score below −2 in children or BMI below 18.5 kg/m² in adults; normal weight as a z score from −2 to 1 in children or BMI 18.5–24.9 kg/m² in adults; and overweight or obesity as a z score above 1 in children or BMI of at least 25 kg/m² in adults. Effect modification was assessed using treatment-by-covariate interaction terms, with a P value below 0.05 considered evidence of interaction.

To evaluate the robustness of the findings, sensitivity analyses were performed by excluding trials at high risk of bias, participants receiving TPT after randomisation, participants diagnosed with incident TB disease within 90 days of initiating study medication, or those with <80% adherence to study medication.

## Results

### Study Selection and Individual Participant Data Obtained

The study selection process is illustrated in Figure 1. Our search identified 123 unique studies that were screened for eligibility, of which four^16–19^ fulfilled eligibility criteria. Studies that were excluded after full-text review are listed, with reasons, in Table S1. IPD were sought from four trials which included 14,653 participants. Data from 34 participants in 1 study^16^ were subsequently excluded from this meta-analysis on the grounds of test-positivity for TBI at baseline. All 14,619 included participants contributed data to co-primary outcomes of incident TBI and/or incident TB disease. Data on serious adverse events were available for 14,533 participants (3 trials). Data on hypercalcaemia were available for 14,533 participants (3 trials) and on renal stones for 10,533 participants (2 trials).

**Figure 1:**
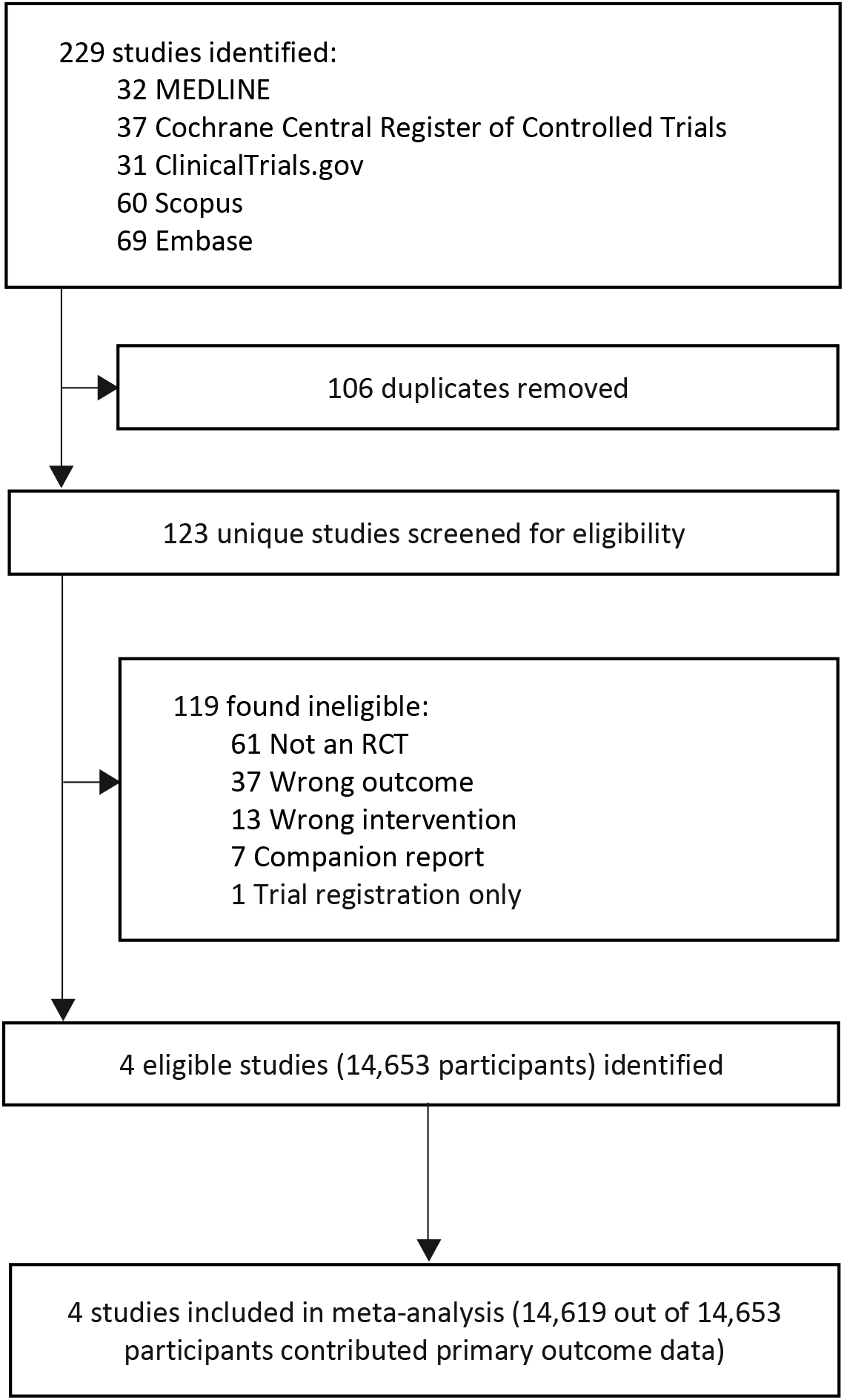
PRISMA Diagram of Study Selection

### Study and Participant Characteristics

The four included trials were conducted in three countries across two continents (Table 1). Three trials enrolled healthy, IGRA-negative school-aged children in Mongolia and South Africa, whereas one enrolled adults living with HIV in Tanzania who were initiating antiretroviral therapy. All were two-arm, parallel-group, placebo-controlled trials investigating oral vitamin D₃ supplementation. Intervention regimens comprised 800 IU daily for 6 months, 14,000 IU weekly for 3 years, 50,000 IU weekly for 1 month followed by 2,000 IU daily for 11 months, or 10,000 IU weekly for 3 years. Follow-up durations ranged from 6 months to 3 years.

**Table 1:** Characteristics of trials and participants included in individual participant data meta-analysis

| ***Study first author & year*** | **Ganmaa 2012<sup>16</sup>** | **Ganmaa 2020<sup>18</sup>** | **Sudfeld 2020<sup>17</sup>** | **Middelkoop 2023<sup>*21*</sup>** |
|---|---|---|---|---|
| ***Setting*** | Mongolia | Mongolia | Tanzania | South Africa |
| ***Participants*** | Healthy school-age children | Healthy, IGRA-negative school aged children | HIV-infected adults initiating ART | Healthy, IGRA-negative school-age children |
| ***Dosing regimen, intervention arm*** | 800 IU<sup>*</sup> oral vitamin D<sub>3</sub> daily for 6 months | 14,000 IU oral vitamin D<sub>3</sub> weekly for 3 years | 50,000 IU oral vitamin D<sub>3</sub> weekly for 1 month, then 2,000 IU daily oral vitamin D<sub>3</sub> for 11 months | 10,000 IU oral vitamin D<sub>3</sub> weekly for 3 years |
| ***Type of control*** | Placebo | Placebo | Placebo | Placebo |
| ***N included / N randomised*** | 86/120<sup>†</sup> | 8851/8851 | 4000/4000 | 1682/1682 |
| ***Intervention: Control*** | 45:41 | 4418:4433 | 2001:1999 | 829:853 |
| ***Mean age, years: included (s.d.) [range]*** | 13.9 (1.2) [12.0 – 16.0] | 9.4 (1.6) [5.8 – 14.2] | 38.7 (9.9) [18.0 – 76.8] | 8.9 (1.4) [5.8 – 11.9] |
| ***Female: Male*** | 42:44 | 4366:4485 | 2735:1265 | 880:802 |
| ***Proportion HIV-infected at baseline*** | Not tested | Not tested | 4000/4000 (100.0%) | 0/1682 (0.0%) |
| ***25(OH)D assay*** | CLIA (LIAISON) | EIA (Immunodiagnostics, UK) | ELFA (VIDAS 25OH Vitamin D total, Biomerieux) | LC-MS/MS |
| ***Vitamin D quality assurance scheme*** | Not reported | DEQAS | Not reported | DEQAS |
| ***Mean baseline 25(OH)D, nmol/L (s.d.) [range]<sup>‡</sup>*** | 15.3 (12.6) [0.0 – 46.9] | 29.6 (10.6) [1.9 – 104.8] | 48.2 (14.7) [0.5 – 75.3] | 71.2 (14.8) [24.9 – 173.6] |
| ***Baseline 25(OH)D <25 nmol/L (%)*** | 70/86 (81.4) | 2819/8846 (31.9) | 288/3998 (7.2) | 1/1364 (0.1) |
| ***Baseline 25(OH)D 25-49.99 nmol/L (%)*** | 16/86 (18.6) | 5636/8846 (63.7) | 1767/3998 (44.2) | 74/1364 (5.4) |
| ***Duration of follow-up*** | 6 months | 3 years | 12 months | 3 years |
| ***TB infection outcome, case definition*** | TST conversion | Positive IGRA test result (interferon-γ level ≥ 0.35 IU/mL) | Not investigated | Positive IGRA test result (interferon-γ level ≥ 0.35 IU/mL) |
| ***TB disease outcome, case definition*** | Not investigated | Positive sputum smear test or clinical diagnosis | Positive sputum smear test, GeneXpert<sup>§</sup>, or clinical diagnosis | Clinical diagnosis (referred for investigation based on symptoms) |

**Footnotes:** \*40,000 international units (IU) vitamin D<sub>3</sub> = 1 mg, †Ganmaa 2012 included a mixed population of TST-positive and TST-negative participants at baseline. N=34 TST-positive participants were excluded from our analysis, ‡25(OH)D concentrations reported in ng/ml were converted to nmol/L by multiplying by 2.496. Seasonally adjusted levels were calculated for Sudfeld 2020. Ganmaa 2020 and Middelkoop 2023 levels were provided seasonally adjusted. Seasonal adjustment was not possible for Ganmaa 2012. Baseline 25(OH)D concentrations were missing for 325 participants: 5 in Ganmaa 2020, 2 in Sudfeld 2020, and 318 in Middelkoop 2023. §GeneXpert MTB/RIF became available at three of the four Sudfeld 2020 study clinics during the trial, where it was used to diagnose TB in smear-negative participants. **Abbreviations:** 25(OH)D, 25-hydroxyvitamin D, ART, antiretroviral therapy, LC-MS/MS, liquid chromatography tandem-mass spectrometry, CLIA, chemiluminescence immunoassay, EIA, enzyme immunoassay, ELFA, enzyme-linked fluorescent assay, IGRA, interferon-gamma release assay. TST, tuberculin skin test.

IPD were available for 14,619 participants, of whom 8,023 (54.9%) were female. Participants ranged in age from 5.8 to 76.8 years. Baseline serum 25-hydroxyvitamin D [25(OH)D] concentrations were available for 14,294 of 14,619 participants (trial mean concentrations 15.3 to 71.2 nmol/L): 3,178 (22.2%) had concentrations below 25 nmol/L, 7,493 (52.4%) had concentrations of 25.0–49.9 nmol/L, and 3,623 (25.3%) had concentrations of 50.0 nmol/L or higher. A total of 7,293 participants (49.9%) were allocated to vitamin D and 7,326 (50.1%) to placebo; baseline characteristics were broadly balanced between treatment groups (Table 1). One trial evaluated incident TBI only, one evaluated incident TB disease only, and two trials evaluated both outcomes. Checks of IPD integrity and consistency identified no material discrepancies between the supplied datasets and the corresponding primary trial reports.

### Risk of Bias within Studies

A Cochrane Risk of Bias assessment was conducted for each of the 4 studies (Table S2) and all studies were determined to be at low risk of bias; therefore our pre-specified sensitivity analysis excluding flagged studies was not deemed to be necessary.

### Overall effect, co-primary outcomes

Overall, vitamin D supplementation did not have a statistically significant effect on either incident TBI or TB disease, in one-stage or two-stage analyses. For incident TBI (9,557 participants in 3 trials), the one-stage adjusted odds ratio (aOR) was 0.97 (95% CI 0.81 to 1.18; P=0.79; Table 2), and the two-stage aOR was 0.91 (95% CI 0.67 to 1.25; I²=46.3%; P for heterogeneity=0.09; Figure 2). For incident TB disease (14,533 participants in 3 trials), the one-stage aOR was 0.78 (95% CI 0.56 to 1.10; P=0.15; Table 3). Two-stage meta-analysis for incident TB disease was not conducted due to trial-level estimates being calculable in only 2/3 studies reporting this outcome. The certainty of evidence was rated as moderate for both co-primary outcomes (Table S3).

**Figure 2:**
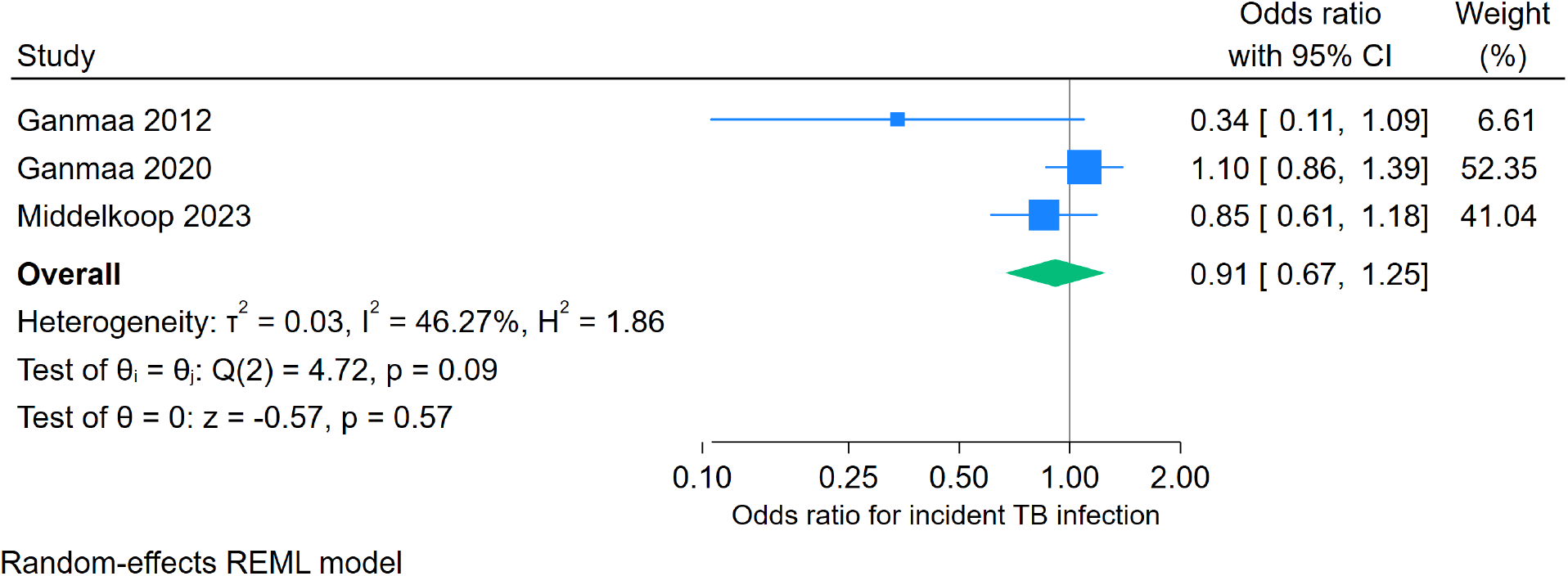
Two-stage individual participant data meta-analysis: **incident tuberculosis infection**, overall.

**Table 2:**
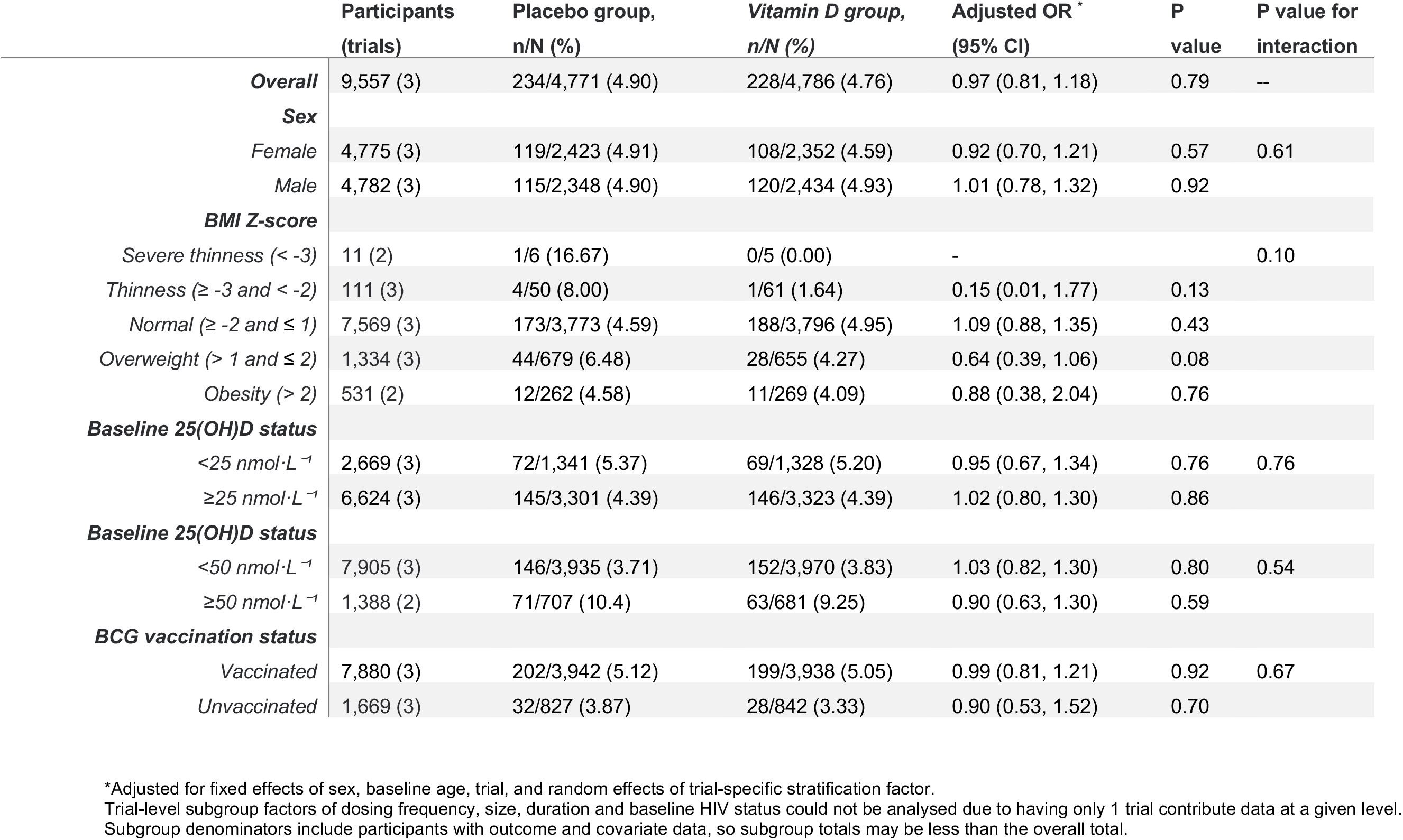
One-stage individual participant data meta-analysis: **incident tuberculosis infection**, overall and by subgroup.

|  | Participants (trials) | Placebo group, n/N (%) | *Vitamin D group, n/N (%)* | Adjusted OR<sup>*</sup> (95% CI) | P value | P value for interaction |
|---|---|---|---|---|---|---|
| ***Overall*** | 9,557 (3) | 234/4,771 (4.90) | 228/4,786 (4.76) | 0.97 (0.81, 1.18) | 0.79 | -- |
| ***Sex*** |  |  |  |  |  |  |
| *Female* | 4,775 (3) | 119/2,423 (4.91) | 108/2,352 (4.59) | 0.92 (0.70, 1.21) | 0.57 | 0.61 |
| *Male* | 4,782 (3) | 115/2,348 (4.90) | 120/2,434 (4.93) | 1.01 (0.78, 1.32) | 0.92 |  |
| ***BMI Z-score*** |  |  |  |  |  |  |
| *Severe thinness (< -3)* | 11 (2) | 1/6 (16.67) | 0/5 (0.00) | - |  | 0.10 |
| *Thinness (≥ -3 and < -2)* | 111 (3) | 4/50 (8.00) | 1/61 (1.64) | 0.15 (0.01, 1.77) | 0.13 |  |
| *Normal (≥ -2 and ≤ 1)* | 7,569 (3) | 173/3,773 (4.59) | 188/3,796 (4.95) | 1.09 (0.88, 1.35) | 0.43 |  |
| *Overweight (> 1 and ≤ 2)* | 1,334 (3) | 44/679 (6.48) | 28/655 (4.27) | 0.64 (0.39, 1.06) | 0.08 |  |
| *Obesity (> 2)* | 531 (2) | 12/262 (4.58) | 11/269 (4.09) | 0.88 (0.38, 2.04) | 0.76 |  |
| ***Baseline 25(OH)D status*** |  |  |  |  |  |  |
| *<25 nmol·L<sup>−1</sup>* | 2,669 (3) | 72/1,341 (5.37) | 69/1,328 (5.20) | 0.95 (0.67, 1.34) | 0.76 | 0.76 |
| *≥25 nmol·L<sup>−1</sup>* | 6,624 (3) | 145/3,301 (4.39) | 146/3,323 (4.39) | 1.02 (0.80, 1.30) | 0.86 |  |
| ***Baseline 25(OH)D status*** |  |  |  |  |  |  |
| *<50 nmol·L<sup>−1</sup>* | 7,905 (3) | 146/3,935 (3.71) | 152/3,970 (3.83) | 1.03 (0.82, 1.30) | 0.80 | 0.54 |
| *≥50 nmol·L<sup>−1</sup>* | 1,388 (2) | 71/707 (10.4) | 63/681 (9.25) | 0.90 (0.63, 1.30) | 0.59 |  |
| ***BCG vaccination status*** |  |  |  |  |  |  |
| *Vaccinated* | 7,880 (3) | 202/3,942 (5.12) | 199/3,938 (5.05) | 0.99 (0.81, 1.21) | 0.92 | 0.67 |
| *Unvaccinated* | 1,669 (3) | 32/827 (3.87) | 28/842 (3.33) | 0.90 (0.53, 1.52) | 0.70 |  |

\*Adjusted for fixed effects of sex, baseline age, trial, and random effects of trial-specific stratification factor.
Trial-level subgroup factors of dosing frequency, size, duration and baseline HIV status could not be analysed due to having only 1 trial contribute data at a given level.
Subgroup denominators include participants with outcome and covariate data, so subgroup totals may be less than the overall total.

**Table 3:** One-stage individual participant data meta-analysis: **incident tuberculosis disease**, overall and by subgroups.

|  | **Participants (trials)** | **Placebo group, n/N (%)** | ***Vitamin D group, n/N (%)*** | **Adjusted OR<sup>\*</sup> (95% CI)** | **P value** | **P value for interaction** |
|---|---|---|---|---|---|---|
| ***Overall*** | 14,533 (3) | 80/7,285 (1.10) | 63/7,248 (0.87) | 0.78 (0.56, 1.10) | 0.15 | -- |
| ***Sex*** |  |  |  |  |  |  |
| *Female* | 7,981 (3) | 47/4,035 (1.16) | 28/3,946 (0.71) | 0.59 (0.37, 0.95) | 0.030 | 0.09 |
| *Male* | 6,552 (3) | 33/3,250 (1.02) | 35/3,302 (1.06) | 1.05 (0.65, 1.71) | 0.84 |  |
| ***BMI (composite)*** |  |  |  |  |  |  |
| *Underweight*<br>*(< -2 z-score children & < 18.5 adults)* | 978 (3) | 24/463 (5.18) | 17/515 (3.30) | 0.64 (0.34, 1.20) | 0.17 | 0.43 |
| *Normal*<br>*(≥ -2 and ≤ 1 z-score children & 18.5-24.9 adults)* | 10,304 (3) | 37/5,172 (0.72) | 35/5,132 (0.68) | 0.96 (0.59, 1.55) | 0.86 |  |
| *Overweight*<br>*(> 1 z-score children & ≥25 adults)* | 3,248 (3) | 19/1,649 (1.15) | 11/1,599 (0.69) | 0.59 (0.29, 1.19) | 0.14 |  |
| ***Baseline 25(OH)D status*** |  |  |  |  |  |  |
| *<25 nmol·L<sup>−1</sup>* | 3,108 (3) | 15/1,593 (0.94) | 13/1,515 (0.86) | 1.01 (0.45, 2.30) | 0.97 | 0.48 |
| *≥25 nmol·L<sup>−1</sup>* | 11,100 (3) | 65/5,528 (1.18) | 50/5,572 (0.90) | 0.75 (0.51, 1.09) | 0.13 |  |
| ***Baseline 25(OH)D status*** |  |  |  |  |  |  |
| *<50 nmol·L<sup>−1</sup>* | 10,585 (3) | 43/5,287 (0.81) | 39/5,298 (0.74) | 0.88 (0.57, 1.37) | 0.58 | 0.39 |
| *≥50 nmol·L<sup>−1</sup>* | 3,623 (3) | 37/1,834 (2.02) | 24/1,789 (1.34) | 0.66 (0.39, 1.11) | 0.11 |  |
| ***BCG vaccination status*** |  |  |  |  |  |  |
| *Vaccinated* | 8,725 (2) | 13/4,391 (0.30) | 10/4334 (0.23) | 0.78 (0.34, 1.79) | 0.56 | 0.63 |
| *Unvaccinated* | 1,808 (2) | 3/895 (0.34) | 3/913 (0.33) | 0.98 (0.40, 2.39) | 0.97 |  |

\*Adjusted for sex, baseline age and trial (fixed effects), with cluster-robust standard errors to account for trial-specific stratification factors.
Trial-level subgroup factors of dosing frequency, size, duration and baseline HIV status could not be analysed due to having only 1 trial contribute data at a given level.
Subgroup denominators include participants with outcome and covariate data, so subgroup totals may be less than the overall total. BCG vaccination status was not available for Sudfeld 2020.

### Sub-group analyses, co-primary outcomes

Results of sub-group analyses by participant-level characteristics from the IPD are presented in Tables 2 and 3. There was no evidence that the effect of vitamin D supplementation on risk of incident TBI differed according to participant sex, baseline BMI, baseline vitamin D status or baseline BCG vaccination status. Similarly, there was no evidence that the effect of vitamin D supplementation on risk of incident TB disease differed according to any of the subgroup factors examined (p-values for interaction >0.05).

### Secondary outcomes

Secondary outcome data were available from three trials comprising 14,533 participants (Table 4). There was no evidence that vitamin D affected the risk of serious adverse events of any cause (aOR 0.95, 95% CI 0.83 to 1.08; P=0.41), non-fatal serious adverse events (aOR 0.89, 0.76 to 1.04; P=0.15) or death from any cause (aOR 1.03, 0.84 to 1.25; P=0.81). Deaths due to tuberculosis (46 with placebo vs 45 with vitamin D, all in the Tanzanian trial) and hypercalcaemia were too few for adjusted pooled analysis, and no renal stones were reported.

**Table 4:**
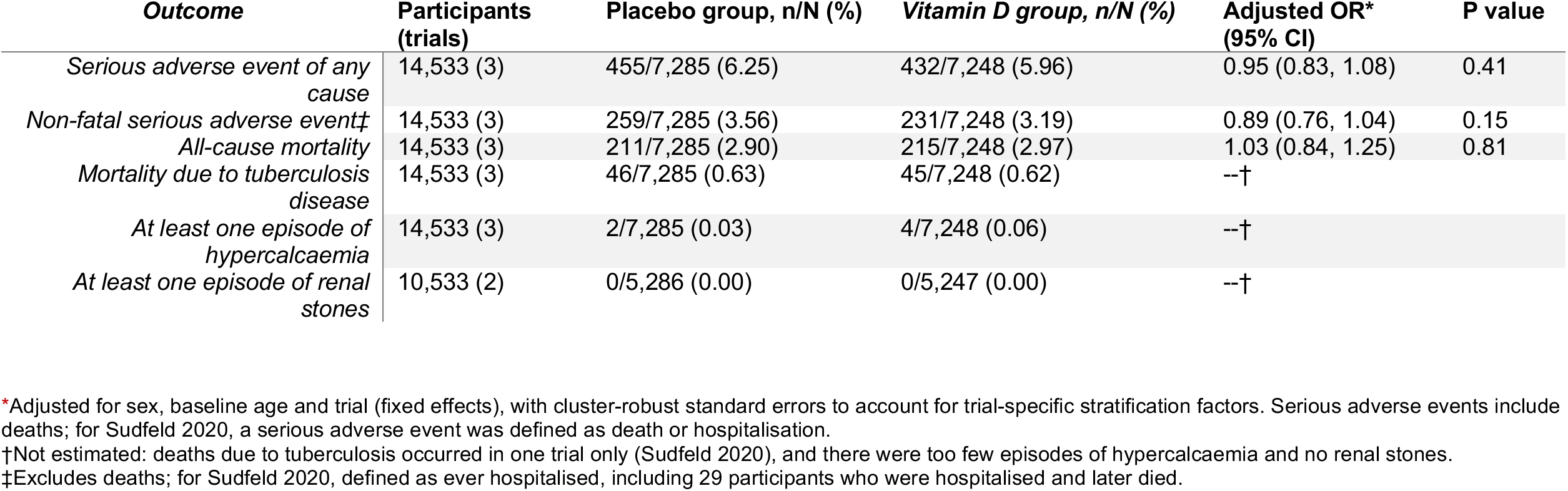
One-stage individual participant data meta-analysis: **secondary outcomes**.

### Sensitivity Analyses

Sensitivity analyses (Table S4) were conducted where possible using one-stage IPD meta- analysis to evaluate the robustness of findings for the co-primary outcomes. For incident TB disease, effect estimates remained non-significant after excluding participants who initiated TPT during follow-up; who were diagnosed with TB disease within 90 days of receiving their first dose of study medication; and who received less than 80% of scheduled study medication doses (P>0.05). For incident TBI, restriction to participants who received at least 80% of scheduled study medication, also produced a result consistent with the primary analysis (P>0.05).

## Discussion

To our knowledge, this is the first IPD meta-analysis of randomised controlled trials evaluating vitamin D supplementation for the primary prevention of TBI and TB disease. In four eligible placebo-controlled trials contributing data from 14,619 participants, vitamin D supplementation did not reduce the overall risk of incident TBI, as indicated by TST or IGRA conversion, or incident TB disease. The effect estimate for incident TBI was relatively precise (adjusted odds ratio [aOR] 0.97, 95% CI 0.81–1.18), making a large protective effect unlikely, although a modest reduction of up to about 19% in the odds of TBI cannot be excluded. By contrast, the estimate for incident TB disease was less precise (aOR 0.78, 95% CI 0.56– 1.10): although compatible with no effect, its confidence interval also included a potentially clinically important up to 44% reduction in odds. We found no convincing evidence that treatment effects varied by sex, body mass index, baseline circulating 25(OH)D concentration, or BCG vaccination status. Trial-level effect modifiers, including dosing frequency, intervention duration, trial size, and baseline HIV status, could not be evaluated because too few trials contributed data at each level. Certainty of evidence was rated as moderate for both co-primary outcomes (Table S3).

Our findings are consistent with the predominantly null results of the contributing trials and with the inconclusive pooled estimates of two previous aggregate-data meta-analyses,^20,21^ but pooling IPD substantially increased statistical power and precision and enabled treatment effects to be investigated across consistently defined participant-level subgroups. This is the principal advantage of IPD over aggregate-data meta-analysis: in particular, we could examine effects among participants with profound vitamin D deficiency (baseline 25(OH)D <25 nmol/L; n=2,669 for TBI and n=3,108 for TB disease), in whom benefit is most biologically plausible, and found no evidence of benefit for either outcome (TBI aOR 0.95, 95% CI 0.67–1.34; TB disease aOR 1.01, 95% CI 0.45–2.30). The absence of an observed benefit contrasts with laboratory evidence that vitamin D metabolites augment antimycobacterial responses in Mtb-infected macrophages, and epidemiologic evidence from observational studies showing consistent independent associations between lower vitamin D status and increased risk of TBI and TB disease.^14,15^ Several factors might explain this disconnect: 1) reductionist cell culture experiments might not have captured the complexity of host-pathogen interactions *in vivo*; 2) associations between low vitamin D status and TB risk in observational studies might be attributable to reverse causation and residual or unmeasured confounding, and 3) publication or citation bias might have contributed to overestimation of the strength of the preclinical and early clinical evidence. Conversely, limitations of the available phase 3 trials - including under-representation of people with low baseline vitamin D status, intermittent dosing schedules, contamination bias arising from off-trial supplement use, and biological redundancy of the vitamin D pathway for antimicrobial defence might have attenuated or extinguished a genuine protective signal.^27^ In the Tanzanian trial, supplementation began at ART initiation, when a substantial proportion of incident TB may represent prevalent subclinical disease unmasked by immune reconstitution, for which supplementation starting at the same time may have come too late; consistent with this, 55 of 143 TB disease events occurred within 90 days of starting study medication (Table S4).

Our IPDMA study has several strengths. We obtained IPD from all four eligible trials, harmonised outcome definitions and covariates across datasets, calculated seasonally adjusted baseline vitamin D levels where possible, and included participants from diverse geographical settings and risk groups, comprising healthy school-aged children and adults living with HIV. We used complementary one-stage and, where estimable, two-stage methods; undertook prespecified participant-level subgroup analyses; and found that the principal results were robust in sensitivity analyses excluding participants who initiated TPT, developed TB disease within 90 days of randomisation, or received less than 80% of scheduled study medication. All contributing trials were judged to be at low risk of bias. However, several limitations should be acknowledged. First, only four trials were eligible, and only three contributed to each co-primary outcome. This restricted our power to detect sub- group effects, precluding meaningful assessment of several trial-level modifiers, and making their effects difficult to disentangle because characteristics such as study population, HIV status, setting, dosing regimen, and follow-up duration were partly or wholly collinear with specific trials. It also prevented a two-stage analysis of TB disease because estimable trial- specific effects were available from only two contributing studies. Second, no gold-standard test exists for the detection of asymptomatic TBI. TST and IGRA conversion are immunological surrogate outcomes subject to biological and technical variability, which might indicate recent exposure or sensitisation rather than the establishment of viable, persistent infection.^28^ Similarly, TB disease was diagnosed using trial-specific criteria that included clinical diagnosis without microbiological confirmation (Table S5); non-differential misclassification, particularly of smear-negative TB in people living with HIV, would bias effect estimates towards the null. Notably, in exploratory analyses of the Tanzanian trial, vitamin D reduced the incidence of microbiologically confirmed TB and of high bacillary load (≥2+) sputum smear-positive pulmonary TB.^17^ Third, the trials differed in their populations, baseline vitamin D status, duration of follow-up, outcome ascertainment, and supplementation regimens. In particular, the two co-primary outcomes were informed by different populations: all TBI data came from children in Mongolia and South Africa, whereas 114 of 143 TB disease events (80%) occurred in the Tanzanian trial of adults living with HIV, so the pooled estimates may not generalise to other groups at high risk of TB, such as HIV-negative adults or household contacts of people with TB. Daily-equivalent doses ranged from 800 IU (Ganmaa 2012) to approximately 2,000 IU (Ganmaa 2020, and Sudfeld 2020 after a 4-week weekly loading phase), with approximately 1,400 IU in Middelkoop 2023. All regimens raised 25(OH)D concentrations relative to placebo: mean end-of-trial concentrations were 77 vs 27 nmol/L in Ganmaa 2020 and 104 vs 65 nmol/L in Middelkoop 2023, and concentrations were significantly higher with vitamin D throughout follow-up in Sudfeld 2020, whereas 800 IU/day raised mean concentrations only from 17 to 50 nmol/L in Ganmaa 2012, with 60% of children remaining below 50 nmol/L.^16–19^ Inadequate dosing is therefore an unlikely explanation for the null findings of the three larger trials. Three of the four regimens included weekly administration, which may precipitate fluctuations in circulating 25(OH)D concentrations that are not seen with daily dosing. Such fluctuations may induce counter-regulatory hydroxylase activity or provide inadequate daily availability of short-lived ‘parent’ vitamin D_3_ (cholecalciferol), potentially diminishing extra-renal or intracrine responses; therefore, findings arising from use of these regimens might not reflect effects of regular daily supplementation.^29,30^ Baseline 25(OH)D concentrations could not be seasonally adjusted for one small trial (Ganmaa 2012; n=86), although this is unlikely to have materially affected subgroup classification. Finally, incident TB disease was uncommon, limiting precision for this outcome overall and particularly in subgroup analyses. The absence of statistically significant interactions for active TB should consequently not be interpreted as definitive evidence that treatment effects are identical across all subgroups.

Taken together, current evidence does not support routine vitamin D supplementation solely for the prevention of TBI or disease in populations similar to those studied. The relatively precise result for TBI makes a large overall effect unlikely, although a modest reduction of up to about 19% in the odds of TBI cannot be excluded; for incident TB disease, the confidence interval does not rule out a potentially clinically important benefit. Further trials of similar intermittent regimens in schoolchildren are unlikely to change conclusions for TBI; for TB disease, however, additional trials in high-risk populations would improve precision and could confirm or exclude a clinically important benefit. Such trials should focus on populations at high risk of TB disease in whom benefit remains most plausible, such as people living with HIV, household contacts of people with TB, and people with profound vitamin D deficiency (baseline 25(OH)D <25 nmol/L), and should use daily rather than intermittent bolus dosing.

## Data Availability

De-identified individual participant data were provided by the investigators of each contributing trial under data transfer agreements and cannot be shared by the authors; requests for these data should be directed to the investigators of the individual trials. The study protocol (PROSPERO CRD420251173446) and Stata analysis code will be available with publication on reasonable request to.

## Acknowledgements

This work was supported by Barts Charity through its funding of the Queen Mary and Barts Health Tuberculosis Centre (grant number G-002700), which provided DAJ’s salary. ARM was supported by the United Kingdom Office for Students. KM was supported by the Desmond Tutu Health Foundation, Gates Medical Research Institute (Gates MRI-TBV02-301; Gates MRI-TBD-09-201) and TB Alliance (NC-009). The views expressed are those of the authors and not necessarily those of Barts Charity or the Office for Students. We thank all the people who participated in the primary randomised controlled trials, and the teams who conducted them.

## Transparency Declaration

DAJ and ARM are the manuscript’s guarantors and affirm that the manuscript is an honest, accurate, and transparent account of the study being reported.

## Contributors

ARM initiated the project. DAJ and ARM wrote the study protocol, conducted the literature search, assessed eligibility of studies for inclusion and performed risk of bias assessments. KM, DG, IB, CRS and WWF were all directly involved in the acquisition and supply of individual participant data for the work. DAJ designed and executed statistical analyses, with input from ARM. JDS conducted a statistical review and replication of co-primary analysis results. DAJ and JDS had access to, and verified, the underlying individual participant data. DAJ and ARM wrote the first draft of the report. All authors revised it critically for important intellectual content, gave final approval of the version to be published, and agreed to be accountable for all aspects of the work in ensuring that questions related to the accuracy or integrity of any part of the work were appropriately investigated and resolved.

## Declaration of generative AI and AI-assisted technologies

DAJ used Claude (Claude Opus 5.5; Anthropic) to assist in the literature search process: duplicate entries in the merged lists of studies from the 5 databases were flagged by AI and verified by DAJ to be duplicates. DAJ and ARM reviewed the final list of potentially eligible studies for inclusion in the meta-analysis.

## References

1. World Health Organization. Global tuberculosis report 2025. Geneva: World Health Organization; 2025. ISBN: 978-92-4-011692-4. *World Health Organization* 2025.

2. Houben RM, Dodd PJ. The Global Burden of Latent Tuberculosis Infection: A Re- estimation Using Mathematical Modelling. PLoS Med 2016; 13(10): e1002152.

3. Schwalb A, Dodd PJ, Rickman HM, Ugarte-Gil CA, Horton KC, Houben R. Estimating the global burden of viable Mycobacterium tuberculosis infection: A mathematical modelling study. PLoS Med 2026; 23(2): e1004920.

4. Horsburgh CR, Jr. Priorities for the treatment of latent tuberculosis infection in the United States. N Engl J Med 2004; 350(20): 2060–7.

5. Selwyn PA, Hartel D, Lewis VA, et al. A prospective study of the risk of tuberculosis among intravenous drug users with human immunodeficiency virus infection. N Engl J Med 1989; 320(9): 545–50.

6. Tait DR, Hatherill M, Van Der Meeren O, et al. Final Analysis of a Trial of M72/AS01(E) Vaccine to Prevent Tuberculosis. N Engl J Med 2019; 381(25): 2429–39.

7. Smieja MJ, Marchetti CA, Cook DJ, Smaill FM. Isoniazid for preventing tuberculosis in non-HIV infected persons. Cochrane Database Syst Rev 2000; 1999(2): CD001363.

8. Schmidt AC, Fairlie L, Hellstrom E, et al. BCG Revaccination for the Prevention of Mycobacterium tuberculosis Infection. N Engl J Med 2025; 392(18): 1789–800.

9. Dos Santos PCP, Messina NL, de Oliveira RD, et al. Effect of BCG vaccination against Mycobacterium tuberculosis infection in adult Brazilian health-care workers: a nested clinical trial. Lancet Infect Dis 2024; 24(6): 594–601.

10. Rockett KA, Brookes R, Udalova I, Vidal V, Hill AV, Kwiatkowski D. 1,25- Dihydroxyvitamin D3 induces nitric oxide synthase and suppresses growth of Mycobacterium tuberculosis in a human macrophage-like cell line. Infect Immun 1998; 66(11): 5314–21.

11. Sly LM, Lopez M, Nauseef WM, Reiner NE. 1alpha,25-Dihydroxyvitamin D3-induced monocyte antimycobacterial activity is regulated by phosphatidylinositol 3-kinase and mediated by the NADPH-dependent phagocyte oxidase. J Biol Chem 2001; 276(38): 35482–93.

12. Liu PT, Stenger S, Li H, et al. Toll-like receptor triggering of a vitamin D-mediated human antimicrobial response. Science 2006; 311(5768): 1770-3.

13. Martineau AR, Wilkinson KA, Newton SM, et al. IFN-gamma- and TNF-independent vitamin D-inducible human suppression of mycobacteria: the role of cathelicidin LL-37. J Immunol 2007; 178(11): 7190–8.

14. Nnoaham KE, Clarke A. Low serum vitamin D levels and tuberculosis: a systematic review and meta-analysis. Int J Epidemiol 2008; 37(1): 113–9.

15. Aibana O, Huang CC, Aboud S, et al. Vitamin D status and risk of incident tuberculosis disease: A nested case-control study, systematic review, and individual-participant data meta- analysis. PLoS Med 2019; 16(9): e1002907.

16. Ganmaa D, Giovannucci E, Bloom BR, et al. Vitamin D, tuberculin skin test conversion, and latent tuberculosis in Mongolian school-age children: a randomized, double-blind, placebo- controlled feasibility trial. Am J Clin Nutr 2012; 96(2): 391–6.

17. Sudfeld CR, Mugusi F, Muhihi A, et al. Efficacy of vitamin D(3) supplementation for the prevention of pulmonary tuberculosis and mortality in HIV: a randomised, double-blind, placebo- controlled trial. Lancet HIV 2020; 7(7): e463–e71.

18. Ganmaa D, Uyanga B, Zhou X, et al. Vitamin D Supplements for Prevention of Tuberculosis Infection and Disease. N Engl J Med 2020; 383(4): 359–68.

19. Middelkoop K, Stewart J, Walker N, et al. Vitamin D Supplementation to Prevent Tuberculosis Infection in South African Schoolchildren: Multicentre Phase 3 Double-Blind Randomised Placebo-Controlled Trial (ViDiKids). Int J Infect Dis 2023; 134: 63–70.

20. Meng J, Li X, Xiong Y, Wu Y, Liu P, Gao S. The role of vitamin D in the prevention and treatment of tuberculosis: a meta-analysis of randomized controlled trials. Infection 2025; 53(3): 1129–40.

21. Liu S, Lin T, Pan Y. Vitamin D supplementation for tuberculosis prevention: A meta- analysis. Biomol Biomed 2025; 26(4): 559–71.

22. Stewart LA, Clarke M, Rovers M, et al. Preferred Reporting Items for Systematic Review and Meta-Analyses of individual participant data: the PRISMA-IPD Statement. Jama 2015; 313(16): 1657–65.

23. Sachs MC, Shoben A, Levin GP, et al. Estimating mean annual 25-hydroxyvitamin D concentrations from single measurements: the Multi-Ethnic Study of Atherosclerosis. Am J Clin Nutr 2013; 97(6): 1243–51.

24. Higgins JP, Altman DG, Gotzsche PC, et al. The Cochrane Collaboration’s tool for assessing risk of bias in randomised trials. BMJ 2011; 343: d5928.

25. Guyatt GH, Oxman AD, Vist GE, et al. GRADE: an emerging consensus on rating quality of evidence and strength of recommendations. BMJ 2008; 336(7650): 924–6.

26. Riley RD, Lambert PC, Abo-Zaid G. Meta-analysis of individual participant data: rationale, conduct, and reporting. BMJ 2010; 340: c221.

27. Martineau AR. Vitamin D for tuberculosis or acute respiratory infections: lost in translation? Curr Opin Infect Dis 2026; 39(2): 147–52.

28. Martineau AR, Chandran S, Palukani W, et al. Toward a molecular microbial blood test for tuberculosis infection. Int J Infect Dis 2024; 141S: 106988.

29. Vieth R. How to optimize vitamin D supplementation to prevent cancer, based on cellular adaptation and hydroxylase enzymology. Anticancer Res 2009; 29(9): 3675–84.

30. Hollis BW, Wagner CL. Clinical review: The role of the parent compound vitamin D with respect to metabolism and function: Why clinical dose intervals can affect clinical outcomes. J Clin Endocrinol Metab 2013; 98(12): 4619–28.

