## Supplementary appendix for "Vitamin D supplementation to prevent tuberculosis infection and disease: systematic review and individual participant data meta-analysis of randomised controlled trials"

#### Table of Contents


1 *Search strategies* ... 2

2 *Table S1: Studies excluded after full-text review, with reasons* ... 4

3 *Table S2: Risk of Bias Assessment* ... 5

4 *Table S3: GRADE Summary of Findings Table* ... 6

5 *Table S4: Results of exploratory sensitivity analyses* ... 7

6 *Table S5: Definitions and ascertainment of the co-primary outcomes in the included trials* ... 8

7 *Table S6: PRISMA-IPD checklist* ... 9

#### 1 Search strategies

**MEDLINE (via PubMed)**
#1 ("Vitamin D"[Mesh] OR cholecalciferol[tiab] OR ergocalciferol[tiab] OR calciferol[tiab] OR "vitamin D"[tiab] OR "vitamin D3"[tiab])
#2 ("Tuberculosis"[Mesh] OR tuberculosis[tiab] OR "latent tuberculosis"[tiab] OR "latent TB"[tiab] OR "tuberculosis infection"[tiab] OR "Tuberculosis infection"[tiab] OR "tuberculosis disease"[tiab] OR "Tuberculosis disease"[tiab])
#3 (prevent*[tiab] OR prophylaxis[tiab] OR incident[tiab] OR "primary prevention"[tiab])
#4 (randomized controlled trial[pt] OR randomised[tiab] OR randomized[tiab])
#5 #1 AND #2 AND #3 AND #4
(No publication-date or language limits)

**Embase (Embase.com)**
#1 ('vitamin d'/exp OR cholecalciferol:ti,ab OR ergocalciferol:ti,ab OR calciferol:ti,ab OR 'vitamin d':ti,ab OR 'vitamin d3':ti,ab)
#2 ('tuberculosis'/exp OR tuberculosis:ti,ab OR 'latent tuberculosis':ti,ab OR 'latent tb':ti,ab OR 'tuberculosis infection':ti,ab OR 'tuberculosis disease':ti,ab)
#3 (prevent*:ti,ab OR prophylaxis:ti,ab OR incident:ti,ab OR 'primary prevention':ti,ab)
#4 ('randomized controlled trial'/exp OR randomised:ti,ab OR randomized:ti,ab)
#5 #1 AND #2 AND #3 AND #4
(No publication-date or language limits)

**Scopus**
#1 TITLE-ABS-KEY("Vitamin D" OR cholecalciferol OR ergocalciferol OR calciferol OR "vitamin D3")
#2 TITLE-ABS-KEY(tuberculosis OR "latent tuberculosis" OR "latent TB" OR "tuberculosis infection" OR "tuberculosis disease")
#3 TITLE-ABS-KEY(prevent* OR prophylaxis OR incident OR "primary prevention")
#4 TITLE-ABS-KEY("randomized controlled trial" OR randomised OR randomized OR "randomised controlled trial")
#5 #1 AND #2 AND #3 AND #4
(No publication-date or language limits)

**Cochrane Library + Clinical Trials registry**
#1 ([mh "Vitamin D"]) OR (cholecalciferol:ti,ab,kw OR ergocalciferol:ti,ab,kw OR calciferol:ti,ab,kw OR "vitamin D":ti,ab,kw OR "vitamin D3":ti,ab,kw)
#2 ([mh "Tuberculosis"]) OR (tuberculosis:ti,ab,kw OR "latent tuberculosis":ti,ab,kw OR "latent TB":ti,ab,kw OR "tuberculosis infection":ti,ab,kw OR "tuberculosis disease":ti,ab,kw)
#3 (prevent*:ti,ab,kw OR prophylaxis:ti,ab,kw OR incident:ti,ab,kw OR "primary prevention":ti,ab,kw)
#4 (randomi*:ti,ab,kw OR placebo:ti,ab,kw OR trial:ti,ab,kw)
#5 #1 AND #2 AND #3 AND #4

**ClinicalTrials.gov (advanced search)**

**Condition/Disease:** Tuberculosis OR Latent Tuberculosis OR Tuberculosis Infection OR Tuberculosis Disease
**Other terms:** “vitamin D” OR cholecalciferol OR ergocalciferol
**Study type:** Interventional (Clinical Trial)
**Status:** All
**First-posted:** All dates

#### 2 Table S1: Studies excluded after full-text review, with reasons

| First author, year | Reason for exclusion |
|---|---|
| Dude, 2022 | Ineligible: copy of Middelkoop 2023 |
| Yani, 2018 | Ineligible: unpublished, abstract only |

#### 3 Table S2: Risk of Bias Assessment

| | Ganmaa 2012 | Ganmaa 2020 | Sudfeld 2020 | Middelkoop 2023 |
|---|---|---|---|---|
| **Sequence generation** | ü | ü | ü | ü |
| **Allocation concealment** | ü | ü | ü | ü |
| **Blinding of participants and personnel** | ü | ü | ü | ü |
| **Blinding of outcome assessment** | ü | ü | ü | ü |
| **Incomplete outcome data** | ü | ü | ü | ü |
| **Selective reporting** | ü | ü | ü | ü |
| **Other bias** | ü | ü | ü | ü |

**ü = low risk of bias; ? = unclear risk of bias, N/A = not applicable (unpublished)**

### 4 Table S3: GRADE Summary of Findings Table

**Vitamin $D_3$ compared to placebo for prevention of incident TB infection/disease**

**Population**: healthy school-age children and adults receiving therapy for treatment of HIV
**Setting**: three countries, on two continents (South Africa, Tanzania, Mongolia)
**Intervention**: oral vitamin $D_3$ (cholecalciferol) supplementation
**Comparison**: oral placebo

| Outcomes | **Anticipated absolute effects*** (95% CI) | | Relative effect (95% CI) | № of participants (studies) | Quality of the evidence (GRADE) |
|---|---|---|---|---|---|
| | **Risk with placebo** | **Risk with Vitamin D** | | | |
| Proportion with incident TB infection | 49 per 1,000 | 48 per 1,000 (40 to 57) | OR 0.97 (0.81 to 1.18) | 9557 (3 RCTs) | ⊕⊕⊕ MODERATE |
| Proportion with incident TB disease | 11 per 1,000 | 9 per 1,000 (6 to 12) | OR 0.78 (0.56 to 1.10) | 14533 (3 RCTs) | ⊕⊕⊕ MODERATE |
| Proportion with serious adverse event, any cause | 62 per 1,000 | 59 per 1,000 (52 to 67) | OR 0.95 (0.83 to 1.08) | 14533 (3 RCTs) | ⊕⊕⊕ MODERATE |

***The risk in the intervention group** (and its 95% confidence interval) is based on the assumed risk in the comparison group and the **relative effect** of the intervention (and its 95% CI).

**CI:** Confidence interval; **OR:** Odds ratio

**GRADE Working Group grades of evidence**
**High quality:** We are very confident that the true effect lies close to that of the estimate of the effect
**Moderate quality:** We are moderately confident in the effect estimate: The true effect is likely to be close to the estimate of the effect, but there is a possibility that it is substantially different
**Low quality:** Our confidence in the effect estimate is limited: The true effect may be substantially different from the estimate of the effect
**Very low quality:** We have very little confidence in the effect estimate: The true effect is likely to be substantially different from the estimate of effect

#### 5 Table S4: Results of exploratory sensitivity analyses

| *Outcome, analysis* | Participants (trials) | Placebo group, n/N (%) | Vitamin D group, n/N (%) | Adjusted OR[1] (95% CI) | P value |
|---|---|---|---|---|---|
| *TB Disease, excluding those initiating preventative therapy during follow-up* | 14,471 (3) | 79/7,258 (1.09) | 61/7,213 (0.85) | 0.77 (0.55, 1.08) | 0.13 |
| *TB Disease, excluding those diagnosed within 90 days of first vitamin D dose* | 14,478 (3) | 48/7,253 (0.66) | 40/7,225 (0.55) | 0.83 (0.53, 1.29) | 0.41 |
| *TB Disease, excluding those with <80% adherence to vitamin D dosing regimen* | 11,974 (3) | 57/5,967 (0.96) | 41/6,007 (0.68) | 0.70 (0.45, 1.08) | 0.11 |
| *TB Infection, excluding those with <80% adherence to vitamin D dosing regimen* | 8,674 (2)* | 227/4,325 (5.25) | 218/4,349 (5.01) | 0.96 (0.79, 1.16) | 0.65 |

* Adherence data not available for Ganmaa 2012. Ganmaa 2012 did not assess TB disease, so the TB disease analyses include all three trials that assessed this outcome.

#### 6 Table S5: Definitions and ascertainment of the co-primary outcomes in the included trials

| | Ganmaa 2012 | Ganmaa 2020 | Sudfeld 2020 | Middelkoop 2023 |
|---|---|---|---|---|
| TB infection: test | Tuberculin skin test (Mantoux; 2 TU purified protein derivative) | QuantiFERON-TB Gold In-Tube | Not assessed | QuantiFERON-TB Gold Plus |
| TB infection: definition of incident infection | Induration ≥10 mm at follow-up in a child with induration <10 mm at baseline | Interferon-γ ≥0.35 IU/mL at follow-up in a child with a negative result at screening | Not assessed | Interferon-γ ≥0.35 IU/mL at follow-up in a child with a negative result at baseline |
| TB infection: timing of testing | Baseline and 6 months | Screening and 3 years | Not assessed | Baseline and 3 years |
| TB disease: case finding | Not assessed | Recorded at weekly visits for supervised dosing during school terms; children with QFT conversion at 3 years referred for clinical and radiographic screening | Symptom screen at every study visit (weekly for 3 weeks, then monthly); people with suspected TB gave three sputum samples and had a chest X-ray | Referral for investigation on the basis of symptoms |
| TB disease: case definition | Not assessed | Diagnosis by local clinicians (sputum smear or clinical diagnosis), adjudicated by the trial end-point committee | ≥1 sputum smear positive for acid-fast bacilli, or chest X-ray features suggestive of TB without a positive smear; GeneXpert MTB/RIF was used for smear-negative individuals at three of four clinics once it became available during the trial | Clinical diagnosis |

Sources: published reports of the included trials. QFT, QuantiFERON-TB Gold; TB, tuberculosis; TU, tuberculin units.

### 7 Table S6: PRISMA-IPD checklist

| Section/topic | Item | PRISMA-IPD checklist item (abridged) | Reported in |
|---|---|---|---|
| **Title** | | | |
| Title | 1 | Identify the report as a systematic review and meta-analysis of individual participant data (IPD) | Title page |
| **Abstract** | | | |
| Structured summary | 2 | Structured summary: objectives; eligibility criteria; data sources and date of last search; that IPD were sought; risk-of-bias methods; numbers of studies and participants identified and obtained; summary estimates with CIs and heterogeneity; interpretation; funding; registration | Summary (heterogeneity statistics are in Results: Overall effect, co-primary outcomes) |
| **Introduction** | | | |
| Rationale | 3 | Rationale for the review in the context of what is already known | Introduction, paragraphs 1–3; Research in context |
| Objectives | 4 | Questions addressed (participants, interventions, comparisons, outcomes, study design), including hypotheses about participant-level subgroups | Introduction, final paragraph |
| **Methods** | | | |
| Protocol and registration | 5 | Whether a protocol exists, where it can be accessed, and registration details | Methods: Search strategy and selection criteria, paragraph 1 (PROSPERO CRD420251173446) |
| Eligibility criteria | 6 | Inclusion and exclusion criteria, whether applied at study or participant level, and rationale | Methods: Search strategy and selection criteria, paragraph 2 (participants with the outcome at baseline excluded at participant level) |
| Identifying studies: information sources | 7 | All methods used to identify published and unpublished studies, and date of last search | Methods: Search strategy and selection criteria, paragraph 3; Figure 1 (MEDLINE, Embase, Scopus, CENTRAL and ClinicalTrials.gov, to Oct 29, 2026) |
| Identifying studies: search | 8 | Full electronic search strategy for at least one database, including limits | Appendix, section 1 (strategies for all sources) |
| Study selection processes | 9 | Process for deciding which studies were eligible | Methods: Search strategy and selection criteria, paragraph 3; Figure 1 |
| Data collection processes | 10 | How IPD were requested, collected and managed, including querying data with investigators; handling of any studies without IPD | Methods: Data analysis, paragraph 1 (IPD were obtained for all eligible trials, so no aggregate data were needed) |
| Data items | 11 | How variables were chosen; study-level and participant-level data sought; any standardisation of variables across studies | Methods: Data analysis, paragraphs 2–3 (including 25(OH)D assays and seasonal adjustment); Table 1 |
| IPD integrity | A1 | Aspects of the IPD that were checked, and how | Methods: Data analysis, paragraph 1 |
| Risk of bias assessment in individual studies | 12 | Methods for assessing risk of bias, and any use of IPD checks or risk of bias in the synthesis | Methods: Data analysis (risk of bias and GRADE paragraph) |
| Specification of outcomes and effect measures | 13 | Treatment comparisons; definitions of all outcomes; whether prespecified and primary or secondary; effect measures | Methods: Data analysis (outcome definitions paragraph); Table S5 |
| Synthesis methods | 14 | Meta-analysis methods: one-stage or two-stage approach, models, clustering within studies, fixed or random effects, heterogeneity and missing data | Methods: Data analysis (synthesis paragraphs); analyses used participants with complete outcome and covariate data (Tables 2 and 3, footnotes) |
| Exploration of variation in effects | A2 | Methods for exploring variation in effects, the participant-level characteristics examined as effect modifiers, and whether prespecified | Methods: Data analysis (effect modification paragraph) |
| Risk of bias across studies | 15 | Assessment of risk of bias in the accumulated evidence, including from IPD not obtained | Methods: Data analysis (funnel plots not undertaken with three trials per outcome); IPD were obtained from all eligible trials |
| Additional analyses | 16 | Methods of additional and sensitivity analyses, and whether prespecified | Methods: Data analysis, final paragraph |
| **Results** | | | |
| Study selection and IPD obtained | 17 | Numbers of studies screened, assessed and included, with reasons for exclusion; studies and participants for which IPD were sought and obtained; flow diagram | Results: Study selection and IPD obtained; Figure 1; Table S1 |
| Study characteristics | 18 | Key study and participant characteristics for each study, with citations | Results: Study and participant characteristics; Table 1; Table S5 |

| Section/topic | Item | PRISMA-IPD checklist item (abridged) | Reported in |
|---|---|---|---|
| IPD integrity | A3 | Important issues found when checking the IPD, or a statement that there were none | Results: Study and participant characteristics, final sentence |
| Risk of bias within studies | 19 | Risk of bias assessments, and their bearing on the robustness of conclusions | Results: Risk of bias within studies; Table S2 |
| Results of individual studies | 20 | For each study and main outcome: numbers analysed, summary data by group, and effect estimates with CIs | Figure 2 (incident tuberculosis infection); trial-level estimates for tuberculosis disease were not estimable for all trials (Results: Overall effect, co-primary outcomes) |
| Results of syntheses | 21 | Summary effects with CIs and heterogeneity; numbers of studies, participants and events; interaction estimates for each effect modifier | Results: Overall effect, Sub-group analyses and Secondary outcomes; Tables 2–4; Figure 2 |
| Risk of bias across studies | 22 | Results of any assessment of risk of bias across studies | Results: Overall effect, co-primary outcomes (certainty of evidence); Table S3 |
| Additional analyses | 23 | Results of additional and sensitivity analyses | Results: Sensitivity analyses; Table S4 |
| **Discussion** | | | |
| Summary of evidence | 24 | Main findings and strength of evidence for each main outcome | Discussion, paragraph 1 |
| Strengths and limitations | 25 | Strengths and limitations of the evidence, including the benefits of IPD and any limitations from IPD not obtained | Discussion, paragraph 3 |
| Conclusions | 26 | General interpretation of the findings in the context of other evidence | Discussion, paragraphs 2 and 4; Research in context |
| Implications | A4 | Relevance to key groups, and implications for future research | Discussion, final paragraph; Research in context (Implications of all the available evidence) |
| **Funding** | | | |
| Funding | 27 | Sources of funding and other support (including supply of IPD), and their role | Summary (Funding); Methods: Role of the funding source; Acknowledgements; Contributors |

Checklist items abridged from the PRISMA-IPD statement (Stewart LA, Clarke M, Rovers M, et al. JAMA 2015; 313: 1657–65). Locations refer to sections of the main manuscript unless stated otherwise.
